# Deciphering the Non-Coding Genome in Autism Spectrum Disorders (ASD): A Study of De Novo and Rare Inherited Variants through Targeted Sequencing in Regulatory Regions

**DOI:** 10.1101/2024.10.14.24315434

**Authors:** S Dominguez-Alonso, JR Paul Trotta, J Gonzalez-Peñas, M Fernandez-Prieto, M Parellada, C Arango, A Carracedo, C Rodriguez-Fontenla

**Author notes:** Corresponding author: C Rodriguez-Fontenla, Grupo de Medicina Xenómica, Facultad de Medicina, Universidad de Santiago de Compostela, San Francisco s/n., Santiago de Compostela, 15782, Spain. Contributed equally to this work (A Carracedo, C Rodriguez-Fontenla).

## Abstract

ASD (Autism Spectrum Disorders) are NDDs (Neurodevelopmental Disorders) with complex etiology including multiple genetic and environmental factors. Non-coding mutations contribute to the multifactorial etiology of ASD by influencing gene activity through various regulatory mechanisms. Advances in genomic technologies, such as whole-genome sequencing (WGS) and chromatin interaction studies, have highlighted the role of non-coding regions in ASD genetics. Identifying these non-coding variants enhances our understanding of the underlying complex genetic landscape ASD.

This study aims to analyze the impact of non-coding mutations within regulatory regions in Autism Spectrum Disorder (ASD). The research builds upon a cohort of 360 Spanish ASD trios, from which 200 trios were selected after excluding cases with known copy number variants (CNVs) and whole-exome sequencing (WES) mutations. The selection process intentionally enhanced the sample for undiscovered non-coding risk variants by excluding cases with de novo loss-of-function mutations or large de novo CNVs. To identify regulatory regions of interest, the study employed targeting sequencing of a selection of candidate cis-regulatory elements (cCREs) from ENCODE v2. De novo variation and rare inherited variation were studied using different bioinformatic pipelines and their impact on regulatory activities was assessed using a deep-learning approach (Sei framework). Additional analysis including candidate gene elucidation using ATAC-seq and PLAC-seq data in neuronal cells, variant prioritization, protein-protein interaction (PPI) network, Transcription Factor (TF) enrichment, presence in topologically associated domains (TADs) were also carried out. Sex bias in regulatory variation within ASD was also explored in our analysis.

We discovered that 28% of de novo variants and 25% of inherited variants with high regulatory potential were found in patients with negative results from whole-exome sequencing (WES) and microarray analyses, as assessed by Sei. By integrating PLAC-Seq data, we functionally annotated approximately 80% of de novo variants and 85% of inherited variants. While resources like ENCODE provide valuable insights into genomic regulatory elements, it is crucial to be cautious when prioritizing specific regulatory elements based on initial hypotheses regarding their impact on gene regulation: many sequence classes associated with ASD in this study did not show significant enrichment in any particular cCRE signature. Notably, the most important observation in this study is the implication of a global dysregulation of CTCF suggesting a potential mechanistic impact on the chromatin architecture. In addition,We have found that the most high-impact regulatory variants—whether *de novo* or inherited—are linked to genes not previously associated with ASD. Nevertheless, gene ontology (GO) enrichments indicate that both coding and non-coding variations likely interact within already characterized ASD pathways.

## INTRODUCTION

Autism Spectrum Disorder (ASD) is a complex neurodevelopmental disorder (NDD) characterized by challenges in social interaction, communication, and repetitive behaviors. While significant progress has been made in identifying genetic risk factors for ASD, recent research has highlighted the crucial role of non-coding regions of the genome in its etiology. These non-coding elements, which constitute approximately 99% of the human genome, have emerged as important contributors to the genetic landscape of ASD.

Advances in whole-genome sequencing (WGS) technologies and computational methods have enabled researchers to explore the impact of non-coding mutations on ASD risk. Studies have revealed that ASD probands harbor de novo mutations (DNMs) in non-coding regions that disrupt both transcriptional and post-transcriptional regulation, with significantly higher functional impact compared to those found in unaffected siblings. While there is no question that non-coding variation plays a role in ASD, initial WGS studies were limited in scope to less than 100 parent-child trios^1,2^ and it was only 5 years ago that WGS studies significantly expanded, encompassing larger cohorts ranging from 200 to 500 families^3–6^.

Considering the expectation that non-coding mutations may vary widely in functional impact, with only a small fraction likely to exhibit strong effect sizes, detecting associations has proven challenging^7,8^. Under this rationale, power calculations indicate that identifying such signals would necessitate a very large cohort^7,9^. To overcome this, most published studies have restricted to “candidate” non-coding elements, which involves a priori prediction of which regulatory elements of the non-coding genome are important for disease risk^1,10^. In analogy to candidate gene studies, which have consistently struggled to produce robust and replicable results, the selection of candidate regulatory regions is anticipated to yield similar outcomes, due to the multitude of possible combinations involving annotations, cell types, brain regions, and developmental stages^11^.

Conversely, other studies have demonstrated that the overall contribution of *de novo* non-coding mutations is comparable to that of loss-of-function (LoF) coding mutations and missense mutations, although not all ASD proband will have impactful non-coding variation^12^. However, a major challenge has been the definition of functional elements and the interpretation of mutational effects.

We aim to explore the role of *de novo* and inherited rare and ultra-rare non-coding mutations in ASD using an unbiased methodology, in which candidate non-coding regulatory elements were chosen solely based on their activity in tissues relevant to ASD, specifically the brain and gastrointestinal (GI) tissues, based on the growing evidence of a gut-brain connection in ASD. The selection of active cCRES (cis-Regulatory Elements) in brain and GI tissue was done from ENCODE project, a catalog which includes 926,535 active elements with different regulatory signatures. 200 trios ASD (600 samples) were selected on the basis of previous negative results for microarray or whole-exome sequencing (WES), a widely discussed approach known for significantly enhancing the diagnostic yield in WGS^13^.

Following targeted sequencing of the designated regions, *de novo* and rare inherited mutations within the selected regulatory elements were identifed and prioritized using Sei, a recently developed framework for systematically forecasting sequence regulatory activities (see Material and Methods section). The objective was to analyze the mutational burden of probands not merely based on the count of mutations, which are susceptible to both statistical power challenges and confounding factors, such as the rise in mutation counts with parental age. Instead, the focus was on evaluating the functional impact of mutations, enabling an assessment of their relevance and potential association with ASD.

Moreover, in order to gain a deeper insight into how these variants may influence disease risk, we performed several approaches. To functionally assign a target gene for the variants, PLAC-seq data from neurons, oligodendrocytes and microglia (human brain) previously published by, were selected. This is particularly interesting because gene annotations do not always correspond to the nearby gene to the regulatory region. In addition, gene prioritization (based on previous literature) and biological *in silico* characterization of possible effects were carried out.

In conclusion, this study aims to elucidate novel non-coding mutations in ASD risk, and potentially identifying novel regulatory mechanisms. By focusing on tissue-specific regulatory elements and employing advanced computational methods, our study contributes to ASD research in non-coding regions employing a targeting sequencing focusing solely on active regulatory elements.

## MATERIAL AND METHODS

### 1.1 Sample selection

#### 1.1.1 Cohort description

The analysis described herein builds upon the complete sample set examined in Alonso Gonzalez *et al.*^14^, which explored the biological roles of postzygotic and germinal coding mutations in ASD.

DNA extraction from the Spanish ASD samples, consisting of 360 trios, was performed using the GentraPuregene blood kit (Qiagen Inc., Valencia, CA, USA) from peripheral blood.

Participants from Santiago (n = 136) were recruited from the Complexo Hospitalario Universitario de Santiago de Compostela and Galician ASD organizations. Meanwhile, subjects from Madrid (n = 224) were enrolled through the AMITEA program at the Child and Adolescent Department of Psychiatry, Hospital General Universitario Gregorio Marañón.

Inclusion criteria stipulated that only individuals aged 3 years or older were included in the study.

Enrolled participants received a clinical diagnosis of ASD from trained pediatric neurologists or psychiatrists, following the criteria outlined in both the DSM Fourth Edition Text Revision (DSM-IV-TR) and Fifth Edition (DSM-5). Additionally, when deemed necessary, the Autism Diagnostic Observation Schedule (ADOS) and the Autism Diagnostic Interview-Revised (ADI-R) were administered.

All participants, along with their parents or legal representatives, provided written informed consent, and the study was conducted in accordance with the principles outlined in the Declaration of Helsinki.

Ninety trios from the Spanish cohort were already analyzed by Lim *et al.*^15^. The entire Spanish cohort was included in Satterstrom *et al.*^16^ as part of the Autism Sequencing Consortium (ASC), a large-scale international genomic consortium integrating ASD cohorts and sequencing data from over one hundred investigators. All data generated as part of the ASC were transferred to dbGaP with Study Accession: phs000298.v4.p3.

#### 1.1.2 Sample selection

Out of the 360 trios with complete phenotype information, microarray and exome sequencing data, we selected cases with negative results for copy number variants (CNVs) and WES (mutations classified as benign, likely benign or VOUS (variant of uncertain significance)). By selecting cases on the basis of the absence of *de novo* LoF mutations or large *de novo* CNVs in prior WES and microarray data, we are intentionally enhancing the sample for undiscovered non-coding risk variants.

After this exclusion, individuals with a syndromic form of ASD were also discarded, in order to avoid ascertainment biases.

Following this, samples were randomly selected, leading to a total sample size of 200 trios (100 from Santiago, 100 from Madrid), including data for 39 female probands and 161 male probands.

Among the most prevalent comorbidities in our cohort, the following stand out: 41.5% of our patients exhibit ID, 7% have ADHD, and 6.5% experience epilepsy.

### 1.2 Selection of the regulatory regions of interest

For the selection of the regulatory regions of interest, we employed the candidate cis-regulatory elements (cCREs) Registry from ENCODE^17,18^ (version 2, https://screen-v2.wenglab.org/).

Brain and GI tissues with DNAse-Seq data available were selected, both from adult (n = 14) and embryonic tissue (n = 59). For each tissue, cCREs labeled as “Low-Dnase” were excluded, as they are inactive in the given tissue^17^.

From the total of 926,535 human cCREs, we selected those that exhibit activity in a greater number of the interrogated tissues. Thus, we selected cCREs that were active in 36 or more tissues (n = 85,394 cCREs). (See Additional Supplementary Material)

#### 1.2.1 Targeted sequencing

Selected regions were sequenced at the National Center for Genomic Analysis (CNAG) using the KAPA HyperChoice Target Enrichment custom probes.

Samples with sex discrepancies when compared to reported pedigrees were dropped and replaced, along with all other samples from the same trio. Moreover, samples which failed CNAG’s quality control (DNA <100 ng / critical degradation (genomic quality number (GQN) < 3.3) were also removed and consequently substituted, leaving 71 trios from Santiago and 129 from Madrid.

Sequencing reads were aligned to GRCh38/hg38 using the Burrows-Wheeler Aligner^19^. Single-nucleotide variants (SNVs) and small insertions-deletions (indels) (< 50 bp) were discovered using the Genome Analysis Toolkit (GATK)^20^ HaplotypeCaller package version 3.4 (https://github.com/broadinstitute/gatk).

### 1.3 Data processing

#### 1.3.1 Joint genotyping

Raw results were downloaded as single-sample gVCFs (genomic variant call format). All individual gVCFs were combined into one multisample gVCF using GATK version 4.2.2.0.Then, SNVs and indels were jointly called across all samples producing a final multi-sample VCF file (format VCFv4.2).

#### 1.3.2 Variant quality score recalibration (VQSR)

Variant call accuracy was estimated using the GATK Variant Quality Score Recalibration (VQSR) method, following the recommendations provided in the GATK4 Best Practice Workflow for SNP and Indel calling^21^.

#### 1.3.3 Dataset analysis

A summary of the quality control and data cleaning procedure is depicted in Figure 1.

**Figure 1.**
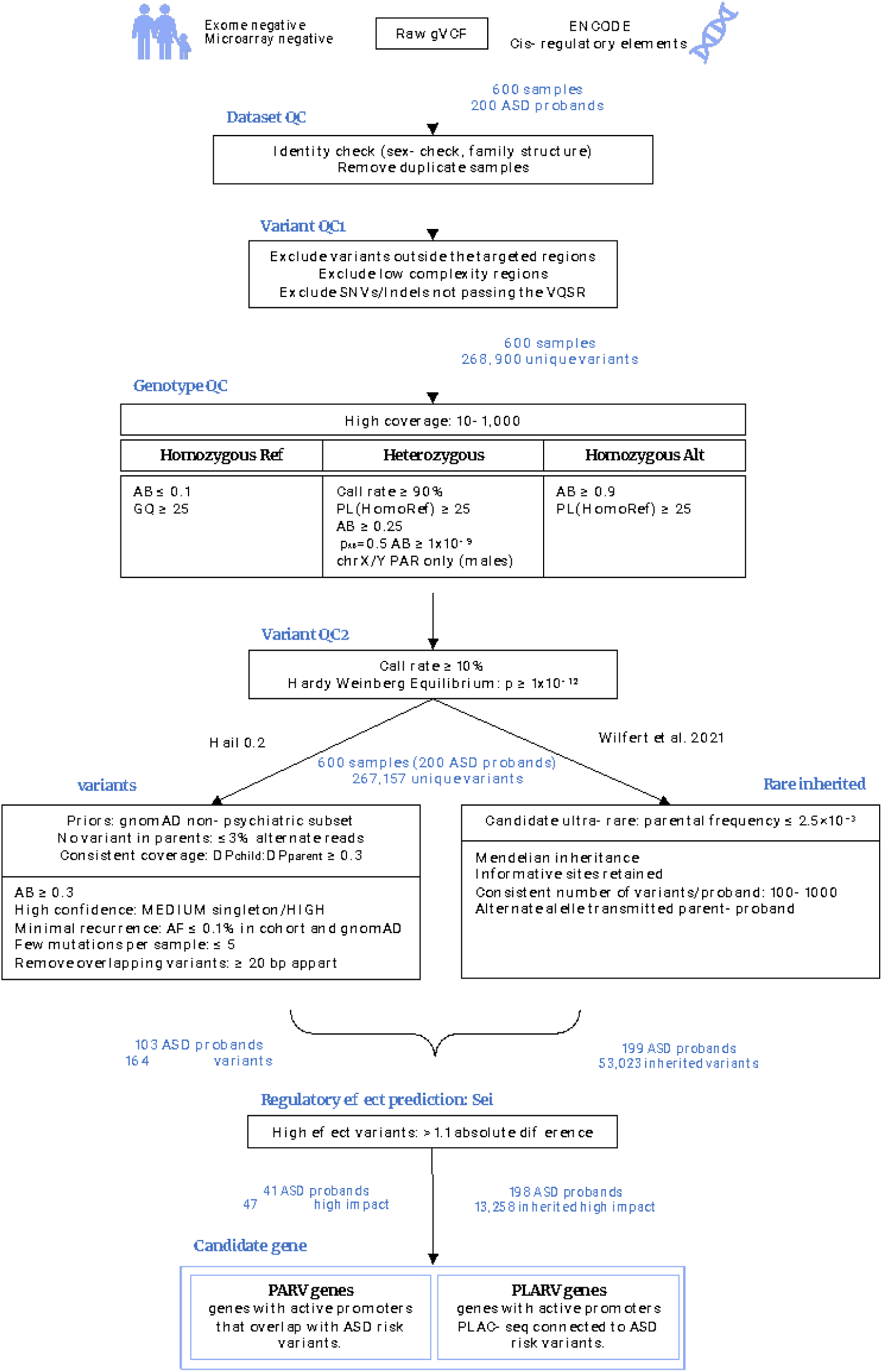
Workflow for variant quality control (QC) and filtering and variant/gene prioritization. *AB, allele balance; AF, allele frequency; bp, base pairs; DP, depth of coverage; GQ, genome quality; PAR, pseudoautosomal regions; PL, phred-scaled likelihood*.

For variant and genotype quality control (QC), as well as optimization of *de novo* variant discovery, we utilized the filtering process established by Satterstrom *et al.*^16^. This process was previously optimized and implemented in an ASD cohort, and we adapted it for our dataset. For inherited variant discovery, we implemented the pipeline defined in Wilfert *et al.*^22^.

##### 1.3.3.1 Dataset Quality Control

The final VCF file, containing SNVs and indels for the 200 trios (600 samples), was loaded into Hail 0.2 (https://hail.is/; https://github.com/hail-is/hail). Then, multiallelic sites were split into biallelic sites (resulting in 719,209 unique variants).

To verify the accuracy of the provided pedigree information, relatedness was assessed between each pair of samples using Hail’s ibd() function. The relatedness values were manually checked, and inferred pedigree structures were compared to reported pedigrees to search for discrepancies. Duplicate samples were identified by using an identity value higher than 0.8 (pi-hat > 0.8) and related samples were identified by using an identity value higher than 0.4 (pi-hat > 0.4)^23^.

No obvious errors in reporting were detected.

##### 1.3.3.2 Variant-level quality control

After filtering the VCF file for the selected coordinates of interest to eliminate off-targets or amplified regions outside the cCREs, low-complexity regions and variants that failed VQSR were excluded. Indels were required to have a variant quality score logs odds (VQSLOD) of ≥ −0.6549, while SNVs were required to possess a VQSLOD of ≥ 5.2528.

Following these steps, a total of 268,900 unique variants remained.

##### 1.3.3.3 Genotype-level quality control

To ensure the quality of genotypes, various filters were implemented during the genotype quality control process.

Initially, we excluded calls with a depth of coverage (DP) < 10 or > 1,000 (as per the distribution of DP values). Moreover, we omitted any call if the GQ was < 25.

For homozygous reference calls, genotypes were then filtered if they had < 90% of the read depth supporting the reference allele.

Homozygous variant calls underwent filtering for genotypes with < 90% of the read depth supporting the alternate allele or a Phred-scaled likelihood (PL) of being homozygous reference < 25.

Heterozygous calls were filtered based on genotypes with < 90% of the read depth supporting either the reference or alternate allele, a PL of being homozygous reference < 25, < 25% of the read depth supporting the alternate allele (*i.e.*, an allele balance (AB) < 0.25), or a probability of the AB (calculated from a binomial distribution centered on 0.5) < 1 x 10^9^.

We additionally omitted any heterozygous call in the X or Y non pseudoautosomal (PAR) regions (in a sample that was imputed as male).

Following the implementation of these filters, variants with a call rate < 10% or a Hardy-Weinberg equilibrium (HWE) p-value <1 x 10^12^ were excluded.

Following the application of these filters and the exclusion of sites that were no longer variants, the dataset comprised 267,157 distinct variants in 200 ASD trios. This dataset was then used as the starting point for the *de novo* and inherited workflows.

#### 1.3.4 *De novo* variant detection

*De novo* variants were identified within the previously described 600-sample dataset.

Variant calling for DNVs utilized the de_novo() function implemented in Hail 0.2. To establish population allele frequencies for all the variants in our dataset, data were sourced from the non-psychiatric subset of gnomAD (https://gnomad.broadinstitute.org/), which includes samples not ascertained for neurological or psychiatric phenotypes (non-neuro subset). Subsequently, these frequencies were utilized as input priors.

Additional criteria for variant calling included the following: (i) homozygous reference genotypes in parents should exhibit no more than 3% of reads supporting the alternate allele, (ii) children’s heterozygous calls were required to have a minimum of 30% of reads supporting the alternate allele, and (iii) the ratio of child DP to parental DP needed to be at least 0.3. This procedure resulted in the identification of 11,311 putative *de novo* variants.

To ensure the quality of *de novo* variants, we retained variants that were classified as high confidence by Hail or were of medium confidence and represented singletons in the dataset (resulting in the inclusion of 10,383 putative *de novo* variants). Subsequently, any variant with an allele frequency exceeding 0.1% across the samples in our dataset (of note, any variant yielded such frequency) or in the non-neuro subset of gnomAD was excluded (leading to the exclusion of 9,739 putative *de novo* variants).

Moreover, samples were excluded if they had more than 5 *de novo* variants (3 samples excluded with 206, 171 and 101 *de novo* variants, respectively), based on z-scores ((data value-mean)/ standard deviation) ε 3.

Additionally, we filtered out variants that were less than 20 base pairs apart from each other to mitigate the likelihood of false positives (resulting in the exclusion of 2 putative *de novo* variants).

After applying these filters, the resultant list of high confidence *de novo* variants included 164 *de novo* variants from 103 probands.

#### 1.3.5 Rare inherited variation

As with *de novo* variation, we used the dataset of 600 samples and 267,157 unique variants described above as a starting point to identify high confidence ultra-rare transmitted variants. For this purpose, we employed the pipeline described by Wilfert *et al.*^22^ (https://github.com/EichlerLab/ultra_rare_transmitted).

Initially, all sites that were heterozygous and observed only once in the parent population (parental frequency ≤ 2.5×10^−3^) were designated as candidate ultra-rare private variants. Subsequently, only variants which did not violate the rules of Mendelian inheritance were kept, resulting in a total of 84,311 variants. Then, only informative sites were retained, leaving 55,903 variants.

After excluding one outlier family with 2,880 transmitted variants (z-score = 13.19), a final set of 53,023 variants was identified in 199 ASD probands.

#### 1.3.6 Regulatory impact prediction

*De novo* and inherited variants were evaluated to determine their impact on regulatory activities using the Sei framework^18^ (https://github.com/FunctionLab/sei-framework).

Sei provides a comprehensive mapping of any given sequence to regulatory activities, classified in 40 distinct sequence classes. It further provides quantitative scores that represent changes in regulatory activities^18^. Within this context, the Sei framework was utilized to acquire sequence class score predictions for the final call set of *de novo*/inherited variants.

For every mutation, we predicted the sequence class scores for both the reference and alternate alleles and computed the sequence class-level variant effect as the predicted scores for the alternate allele subtracting the scores for the reference allele.

For mutations with a strong effect in a different sequence class than the originally assigned sequence class (absolute value higher than the original sequence class by > 1.1 difference) we reassigned the mutation to the sequence class with the strongest effect.

Moreover, variants were annotated in terms of positive effect predictions (increase in the assigned activity of a sequence) or negative effect predictions (decrease in the activity).

These filtering yielded 47 *de novo* variants, in 44 ASD probands with changes in sequence class activity and 13,258 ultra-rare inherited variants in 198 probands.

#### 1.3.7 Candidate gene elucidation

After applying the prioritization criteria for their impact on regulatory activities, we aimed to perform a candidate gene selection. For this purpose, we leveraged brain cell-type specific interactome maps to further confirm our non-coding regulatory regions and pinpoint genes likely to be regulated by these non-coding variants^24^.

In their work, Nott and colleagues^24^ characterized the enhancer-promoter interactome of three different human brain cell types (namely neurons, microglia and oligodendrocytes) *in vivo*. In brief, they defined open regions of chromatin with ATAC-seq and identified active chromatin regions and promoters with ChIP-seq analysis of the H3K27ac and H3K4me3 epigenetic marks, respectively. Following this rationale, they defined putative active promoters (ATAC-seq and H3K4me3 enrichment overlapping H3K27ac peaks within 2,000 bp to a nearest TSS) and enhancers (ATAC-seq plus H3K27ac enrichment in a region outside H3K4me3 peaks) in each cell type. Moreover, they established the relationship between promoters and distal regulatory regions through the application of PLAC-seq. In this method, proximity ligation precedes the enrichment for active promoters using H3K4me3 ChIP-seq. This allowed for the characterization of chromatin loops between active promoters and distal enhancer/superenhancers (*i.e.,* clusters of multiple enhancers).

To better understand the likely functional consequences of our variant call set, we defined putative ASD-risk genes according to whether they exhibit active promoters overlapping with any variant in the call set and/or are linked to active promoters through PLAC-seq defined chromatin interactions.

Initially, the analysis involved determining if each variant intersected with an active promoter in each cell type. Variants that did overlap an active putative promoter were assigned to the corresponding gene, designating these as “genes containing promoters harboring ASD regulatory variants” (from now on, PARV genes).

Subsequently, each variant was extended by 2,500 bp upstream/downstream to correspond with the PLAC-seq bin size of 5,000 bp. These variant-extended regions were then intersected with PLAC-seq bins in each cell type. If a region overlapped one or more PLAC-seq bins distally linked to an active promoter, that region was then assigned to the corresponding “gene containing a promoter PLAC-linked to ASD regulatory variants” (from now on, PLARV genes).

#### 1.3.8 Variant prioritization

We applied additional criteria to prioritize non-coding variants that were already categorized as having a significant regulatory effect, in regard to the putative ASD gene they are regulating, with the aim of considering them as potential causative factors for each affected individual.

For this purpose, we first curated a consensus list of known ASD risk genes from different sources. We collected 291 known ASD genes from the following sources: (i) high-confidence (score = 1) ASD genes collected in SFARI (accessed the 11/2023), (ii) dominant ASD genes reported in recent literature and included in the SPARK genes list (11/2023), (iii) unique genes from recent large ASD genetic studies with an FDR <0.1 (these included references ^16,25,26^), and (iv) autosomal genes with exome-wide significance (p <2.5 × 10^-6^) after combining p-values from enrichment of all *de novo* variants, transmission disequilibrium test (TDT) of rare, inherited LoFs from unaffected parents to affected offspring, and increased rate of LoFs in cases vs population controls^12^.

We then cross-referenced the final dataset of PLARV genes with the custom list of ASD-risk genes and selected those variants where the corresponding PLARV gene was present in the list.

The rationale for excluding PARV genes linked to the variants, as we will delve into later, is that, in certain instances, risk variants were associated with more distant active promoters rather than the nearest gene promoter. A connection with the gene was deemed validated only under functional evidence (*i.e.*, PLAC interactions with PLARV genes) and not genomic localization (variants nearby a PARV gene’s TSS).

#### 1.3.9 Protein-protein interaction (PPI) network

We conducted PPI network analyses and computed network statistics using Cytoscape v.3.7.2 (https://cytoscape.org/).

We employed the multiple protein input option of the STRING database v.11. with default settings, except that we restricted interactions to those of high confidence (0.70). STRING constitutes a biological database encompassing both known and predicted protein-protein interactions. Its implementation is accessible within Cytoscape.

#### 1.3.10 Enrichment analysis: Cluster profiler

ClusterProfiler (https://github.com/YuLab-SMU/clusterProfiler)^27^ was utilized to perform GO over-representation tests.

The package org.Hs.eg.db, provided by Bioconductor, was used as the genome wide annotation for humans.

GO enrichment analysis was performed with specific significance thresholds (p-valueCutoff = 0.01, q-valueCutoff = 0.05) adjusted by the Benjamini-Hochberg procedure. Highly similar GO terms (*e.g.*, > 0.25) were removed by applying the “simplify” function to retain the most representative terms (*i.e.,* the most significant) with parameters: cutoff = 0.25, by = “p.adjust” and select_fun = min.

In order to perform a biological theme comparison between the three cell types, we used the “compareCluster” function, which calculates enriched functional profiles of each gene list and aggregates the results into a single object. For visualization purposes, the “showCategory” parameter, indicating the display of the topmost significant categories, was set to 5.

#### 1.3.11 Transcription factors (TF) enrichment

The analysis of TFs with enriched binding sites in the base positions affected by *de novo* and inherited high regulatory impact variants was conducted using the Enrichment Analysis tool in ChIP-Atlas (https://chip-atlas.org/enrichment_analysis). The specified parameters and input files were as follows: Cell type Class = Neural, Experiment type = ChIP: TFs and others, Threshold for Significance = 50, Dataset A = BED file of the positions of the variants, Dataset B = random permutations of dataset A (x 1000).

#### 1.3.12 Presence in topologically associated domains (TADs)

The interval list of TADs in human tissues and cells (BED format, hg19), was acquired from iPSC-derived neurons. This information was sourced from the Gene Expression Omnibus (GEO) database under the accession number GSE79965 (https://www.ncbi.nlm.nih.gov/geo/query/acc.cgi?acc=GSE79965).

To identify potential associations, prioritized variants were cross-referenced with TAD boundaries. This process aimed to detect instances where multiple variants occurred within the same TAD, providing insights into the spatial organization of genomic elements in the context of these prioritized genetic variations.

## RESULTS

### 2.1 Dataset

We analyzed targeted sequencing data in a cohort of 200 ASD trios, with a ratio of ∼4:1 male probands to females (161 males, 39 females; male-to-female ratio = 4.13), in line with previous estimates^28^.

Based on the evidence of GI and brain tissues playing a role in ASD’s etiology^29^, regions were selected according to their activity in these tissues. This yielded a total of 85,394 cCRES active in 25 brain-tissue samples and 48 GI-tissue samples, spanning a total of 21.35 Mb of the genome (*i.e.,* 0,68% from the total length, 0,70% from the non-coding length)^30^.

The vast majority of selected cCREs corresponded to the ENCODE classification of distal/proximal enhancer-like signature (n = 61,822; 72.40%) whereas promoters were represented to a lesser extent (n = 20,306; 23.78%) (Figure 2), as expected in the basis of the one-to many relationship between promoters and enhancers^24^.

**Figure 2.**
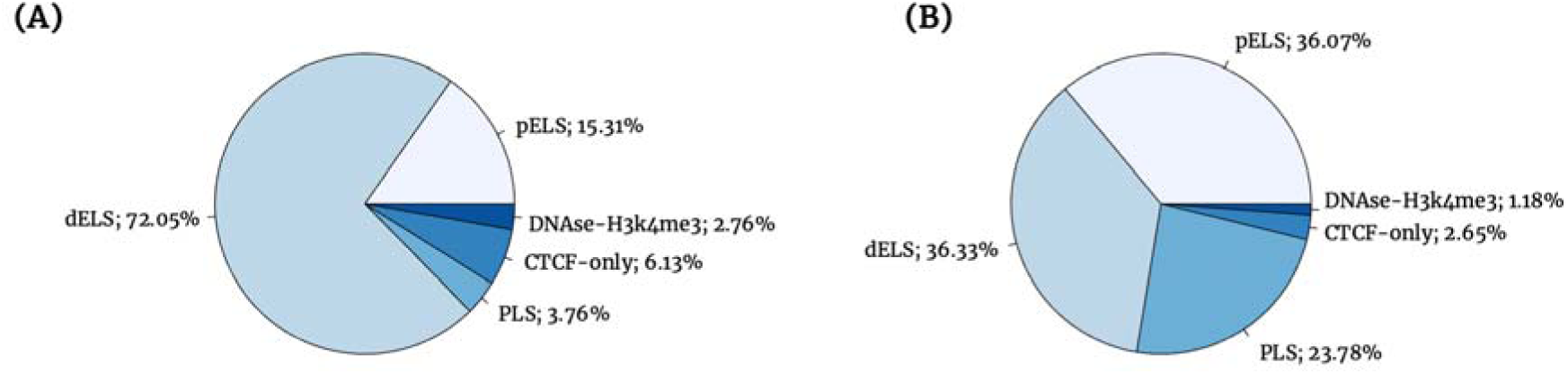
Pie chart depicting the classification of cCREs. Panel A illustrates the distribution of cCREs in the ENCODE registry, while Panel B displays the percentages within our dataset’s selection.

### 2.2 *De novo* variants

From the trio-based data, we identified 164 rare *de novo* variants in the selected candidate regulatory regions (allele frequency ≤ 0.1% in our dataset and in the non-psychiatric of gnomAD), with 51,50% of cases carrying at least one such variant (103 ASD probands; x̅ = 1.59 ± 0.84 variants/child). These variants were present inside an ELS cCRE (both distal and proximal) in 67.08% of the instances, whereas 31.10% were harbored in PLS. The remaining variants were located inside a CTCF-only cCRE (1.22%) and DNAse-H3K4me3 cCREs (0.61%) (Figure 3A).

**Figure 3.**
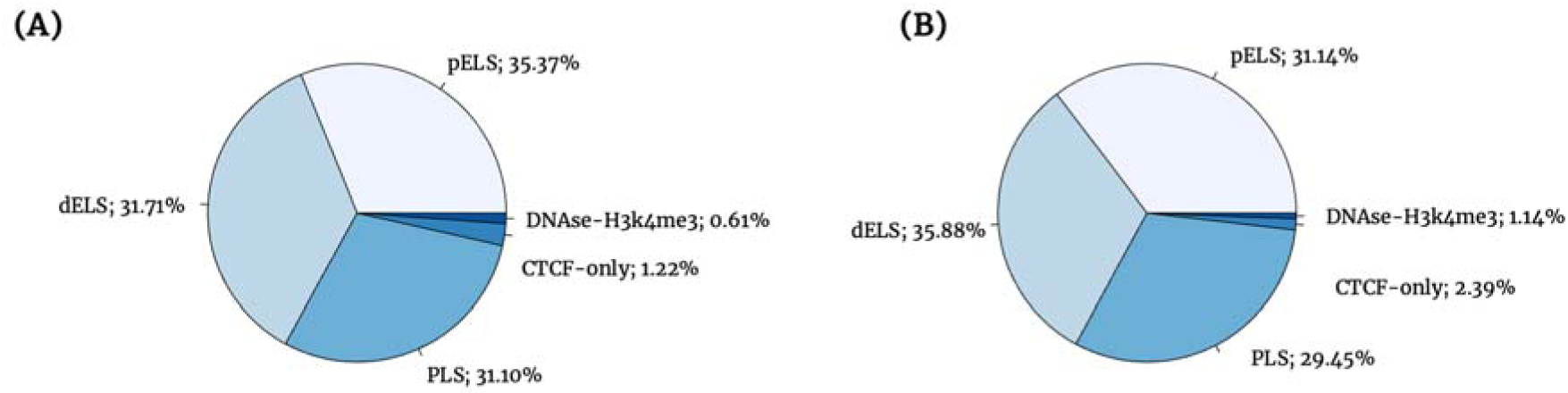
Pie chart representation of *de novo* and inherited variants based on cCREs’ classification. Panel A illustrates the classification for *de novo* variants, while Panel B depicts the classification for inherited variants.

### 2.3 Inherited variants

To study ultra-rare inheritance, we included variants with an allele frequency ≤ 2.5×10^−3^ in the parent population and selected those transmitted to the child. After removing one outlier family with inconsistencies in respect to the average number of mutations per family, 53,023 inherited variants were detected in 199 cases (x̅ = 266.45 ± 68.93 variants/child). These variants were present inside an ELS cCRE in 67.02% of the instances, whereas 29.45% were harbored in PLS cCREs. The remaining variants were located inside a CTCF-only cCRE (2.39%) and DNAse-H3K4me3 cCREs (1.14%) (Figure 3B).

Of note, we observed 26,960 informative sites with a mother-proband inheritance, and 26,063 informative sites with a father-proband inheritance (50.85% *versus* 49.15% of the instances, respectively). The observed differences between these two inheritance patterns were statistically significant (p = 9.97 x 10^-5^; two-sided binomial exact test), consistent with the maternal transmission bias observed for large and small CNVs and inherited private truncating SNVs^1,31–33^. This bias remained significant even when restricting to autosomal chromosomes (p = 7.8 x 10^-3^, two-sided binomial exact test).

Additionally, when restricted to the X chromosome, most of the variants were maternally inherited (97.98% with maternal inheritance pattern *versus* 2.02% with paternal inheritance pattern), as would be expected. Intriguingly, all variants exhibiting an X-linked maternal inheritance pattern were consistently transmitted to an affected son (p = 2.2 x 10^-16^; two-sided binomial exact test) and in any case, to a daughter.

### 2.4 Variant enrichment in cCREs (candidate cis-Regulatory Elements)

To assess potential statistical enrichments or depletions in the number of *de novo* and inherited variants per cCRE category, we conducted two-sample proportions tests.

Comparative analysis with the proportion of each cCRE in the original selection (Figure 2 B) revealed an enrichment of *de novo* and inherited variants in PLS cCREs (p-value for proportion test = 0.02 and < 2.2 x 10^-16^, respectively) (Table 1). Additionally, inherited variants were depleted in CTCF cCREs (p-value = 1.46 x 10^-3^ for the proportion test) and enriched in pELS cCREs (p-value = < 2.2 x 10^-16^ for the proportion test).

**Table 1.** Enrichment of cCREs in *de novo* and inherited variants.

| cCRE | Original selection | De novo | p-value | Inherited | p-value |
| --- | --- | --- | --- | --- | --- |
| dELS | 31,023 | 52 | 0.25 | 19,027 | 0.09 |
| pELS | 30,799 | 58 | 0.92 | 16,510 | $< 2.2 \times 10^{-16}$ |
| PLS | 20,306 | 51 | 0.02 | 15,616 | $< 2.2 \times 10^{-16}$ |
| CTCF | 2,260 | 2 | 0.37 | 1,265 | $1.46 \times 10^{-3}$ |
| DNase-H3K4me3 | 1,006 | 1 | 0.75 | 605 | 0.55 |

### 2.5 Effect of genetic variants in gene regulation

In order to prioritize variants for their potential impact on gene regulatory activities, we used Sei, a new deep learning sequence model that enables the interpretation of genetic variants.

#### 2.5.1 De novo variants

*De novo* variant class scores ranged from 0.07 to 6.68, with 28.66% (n = 47) of the variants yielding absolute differences higher than the stipulated class score > 1.1 difference to consider a variant of high regulatory impact (Supplementary Table 1). These variants were assigned to 10 different regulatory classes in Sei.

Of note, the 47 variants were present in 44 probands, with 3 of them harboring 2 different variants in separate chromosomes (x̅ = 1.07 ± 0.25 *de novo* variants/child).

Half of these variants (53.19%) represented the class CTCF, which demarcates topological loop boundaries^34^. Out of the 25 variants assigned to the regulatory class “CTCF CTCF-Cohesin”, 24 were classified in ENCODE as PLS/dELS/pELS-CTCF bound, and one variant was categorized as CTCF-only.

For the other regulatory classes, variants were inside ENCODE cCREs defined as PLS/dELS/pELS (CTCF bound or not) without following any apparent trend (*i.e*., no regulatory class was significantly correlated with a specific type of cCRE) (data not shown).

#### 2.5.2 Inherited variants

Inherited variant class scores ranged from 0.006 to 41.41, with 25.00% (n = 13,258) of the variants yielding absolute differences higher than > 1.1 (Supplementary Table 2) in 30 different regulatory classes.

The average number of inherited variants was x̅ = 66.96 ± 17.41, and the maximum observed number of inherited variants in a proband was 182.

The CTCF class was the most widely represented in our dataset (40.62% of the variants). In line with the results from *de novo* variants, only 162 variants were present in a CTCF-only cCRE, while the rest were classified in ENCODE as PLS/dELS/pELS/DNase-H3K4me3 CTCF bound or not. Moreover, there were no specific correlations between ENCODE’s cCRE classification restricted to any Sei-assigned class groups, except for CTCF-only cCREs (p-value = 1.79 x 10^-6^ for the proportion test) (Supplementary Table 3).

#### 2.5.3 Differences in gene regulation for inherited and *de novo* variants

*De novo* variants did not yield any significant differences in terms of positive / negative scores neither for the CTCF category (10 variants gained affinity for CTCF binding, and 15 lost this affinity; p-value = 0.42, two-sided binomial test), nor for the rest (Table 2, Figure 4).

**Figure 4.**
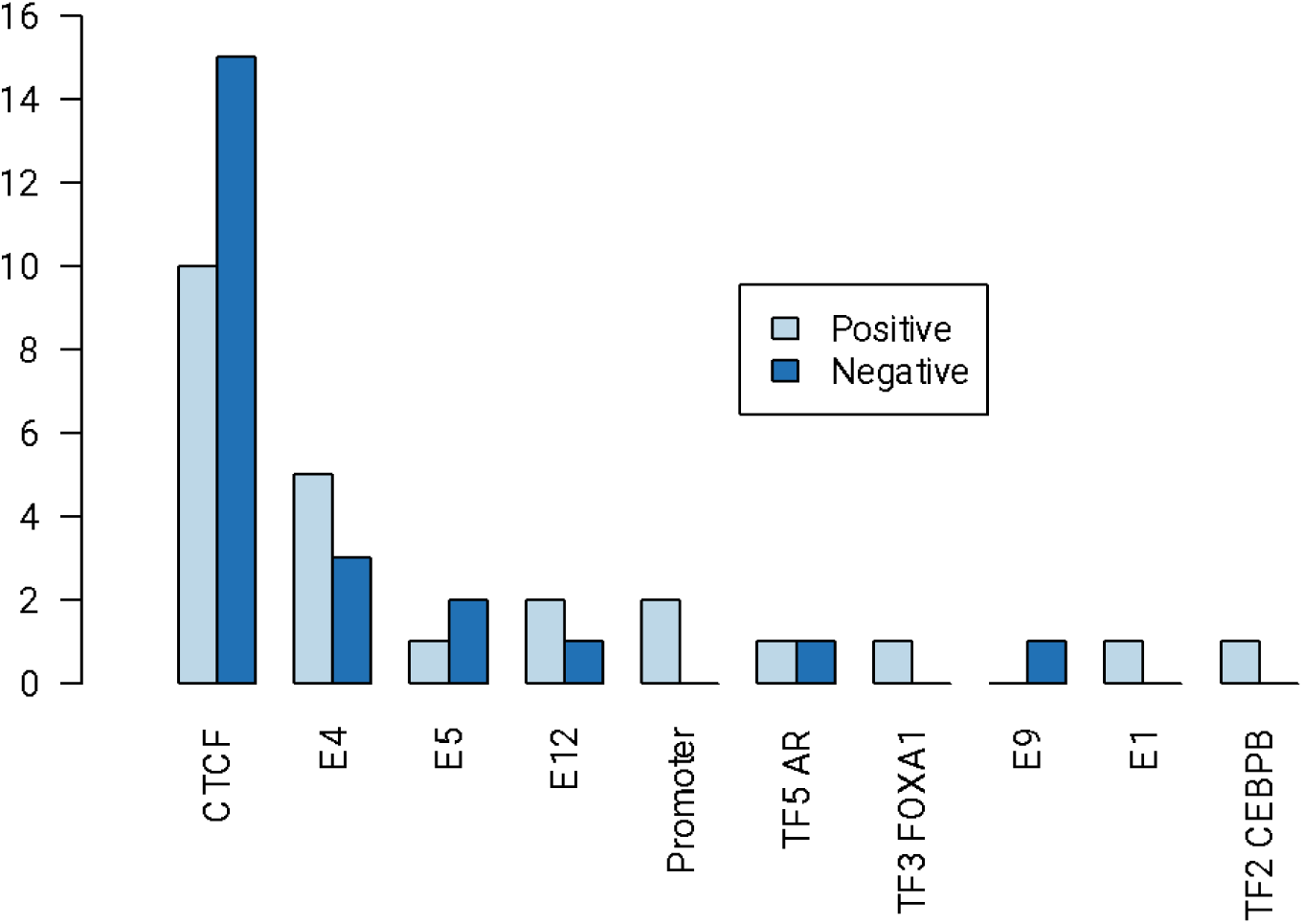
Counts of *de novo* variants assigned to each regulatory class. Regulatory effect scores are categorized into positive and negative scores. No significant differences in positive / negative scores were detected.

**Table 2.** Enrichment of variant scores within each of the regulatory classes, divided in positive and negative effects.

| Regulatory class | *De novo* |  | p-value | Inherited |  | p-value |
| --- | --- | --- | --- | --- | --- | --- |
|  | + | - |  | + | - |  |
| CTCF CTCF Cohesin | 10 | 15 | 0.42 | 2489 | 2896 | **1.55 x 10<sup>-8</sup>** |
| E4 Multi-tissue | 5 | 3 | 0.73 | 1413 | 1930 | **<2.2 x 10<sup>-16</sup>** |
| E5 B cell like | 2 | 1 | 1.00 | 445 | 599 | **1.05 x 10<sup>-6</sup>** |
| E12 Erythroblast like | 2 | 1 | 1.00 | 430 | 588 | **4.11 x 10<sup>-7</sup>** |
| P Promoter | 2 | 0 | 0.50 | 246 | 455 | **1.24 x 10<sup>-15</sup>** |
| TF3 FOXA1 / AR / ESR1 | 1 | 0 | 1.00 | 312 | 378 | **6.64 x 10<sup>-3</sup>** |
| E6 Weak epithelial | - | - | - | 97 | 107 | 0.53 |
| TF5 AR | 1 | 1 | 1.00 | 85 | 69 | 0.23 |
| L5 Low signal | - | - | - | 74 | 54 | 0.09 |
| E7 Monocyte / Macrophage | - | - | - | 65 | 62 | 0.86 |
| E11 T cell | - | - | - | 44 | 39 | 0.66 |
| E9 Liver / Intestine | 0 | 1 | 1.00 | 15 | 44 | **1.02 x 10<sup>-4</sup>** |
| PC4 Polycomb / Bivalent stem cell Enhancer | - | - | - | 29 | 36 | 0.46 |
| E1 Stem cell | 1 | 0 | 1.00 | 28 | 33 | 0.61 |
| TF2 CEBPB | 1 | 0 | 1.00 | 20 | 26 | 0.46 |
| E2 Multi-tissue | - | - | - | 19 | 23 | 0.64 |
| TF4 OTX2 | - | - | - | 21 | 12 | 0.16 |
| E10 Brain | - | - | - | 4 | 10 | 0.18 |
| E8 Weak multi-tissue | - | - | - | 4 | 7 | 0.55 |
| TN1 Transcription | - | - | - | 5 | 4 | 1.00 |
| TN3 Transcription | - | - | - | 5 | 3 | 0.73 |
| E3 Brain / Melanocyte | - | - | - | 4 | 4 | 1.00 |
| PC1 Polycomb / Heterochromatin | - | - | - | 4 | 2 | 0.69 |
| HET4 Heterochromatin | - | - | - | 2 | 3 | 1.00 |
| TN4 Transcription | - | - | - | 2 | 3 | 1.00 |
| PC2 Weak Polycomb | - | - | - | 2 | 2 | 1.00 |
| PC3 Polycomb | - | - | - | 2 | 0 | 0.50 |
| HET5 Centromere | - | - | - | 0 | 1 | 1.00 |
| HET1 Heterochromatin | - | - | - | 1 | 0 | 1.00 |
| TN2 Transcription | - | - | - | 0 | 1 | 1.00 |

For inherited variants, however, we found a significant enrichment in negative scores within the CTCF regulatory class (*i.e.*, a significant decrease in the affinity for CTCF; p-value = 1.55 x 10^-8^, one-sided binomial test) (Table 2, Figure 5). Besides the CTCF class, 6 additional regulatory classes yielded a significant enrichment in negative scores (Table 2).

**Figure 5.**
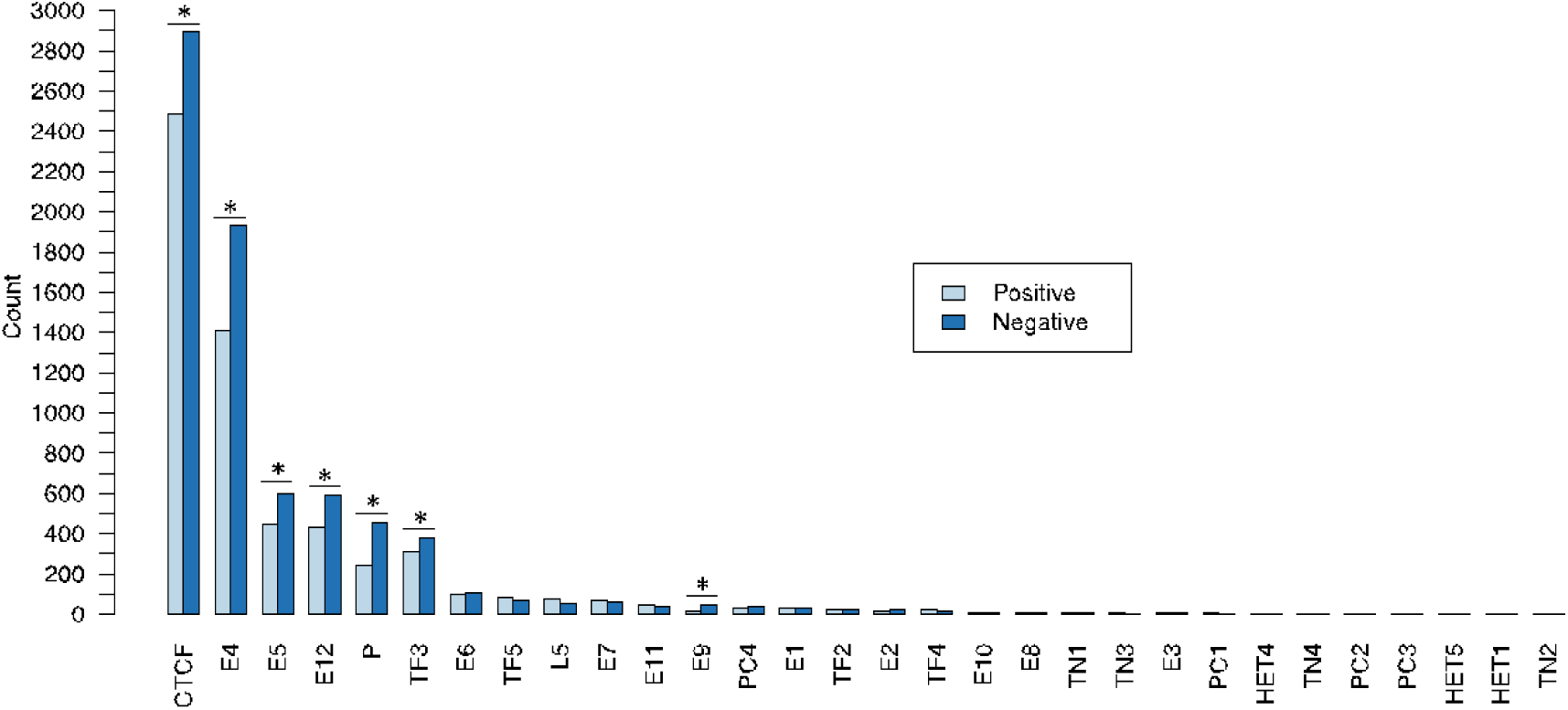
Counts of inherited variants assigned to each regulatory class. Regulatory effect scores are categorized into positive and negative scores. *Significant differences in the number of variants with positive or negative scores (two-sided binomial test).

### 2.6 Candidate gene association

To assess the enrichment of genetic variations linked to complex traits within cell type-specific regulatory regions, Nott and collaborators conducted a linkage disequilibrium score (LDSC) regression analysis of heritability. In their LDSC approach, employing GWAS summary statistics from Grove *et al.*^35^, they revealed that the SNP heritability of ASD was significantly enriched in regulatory elements specific to neurons, particularly in neuronal enhancers. In light of these results, our analysis will predominantly be centered on neuronal-specific regulatory regions.

#### 2.6.1 De novo

From the total of 47 high regulatory impact *de novo* variants, 29 fell inside a putative promoter (61.70% of the instances), all of them corresponding to a different PARV gene (Figure 6).

**Figure 6.**
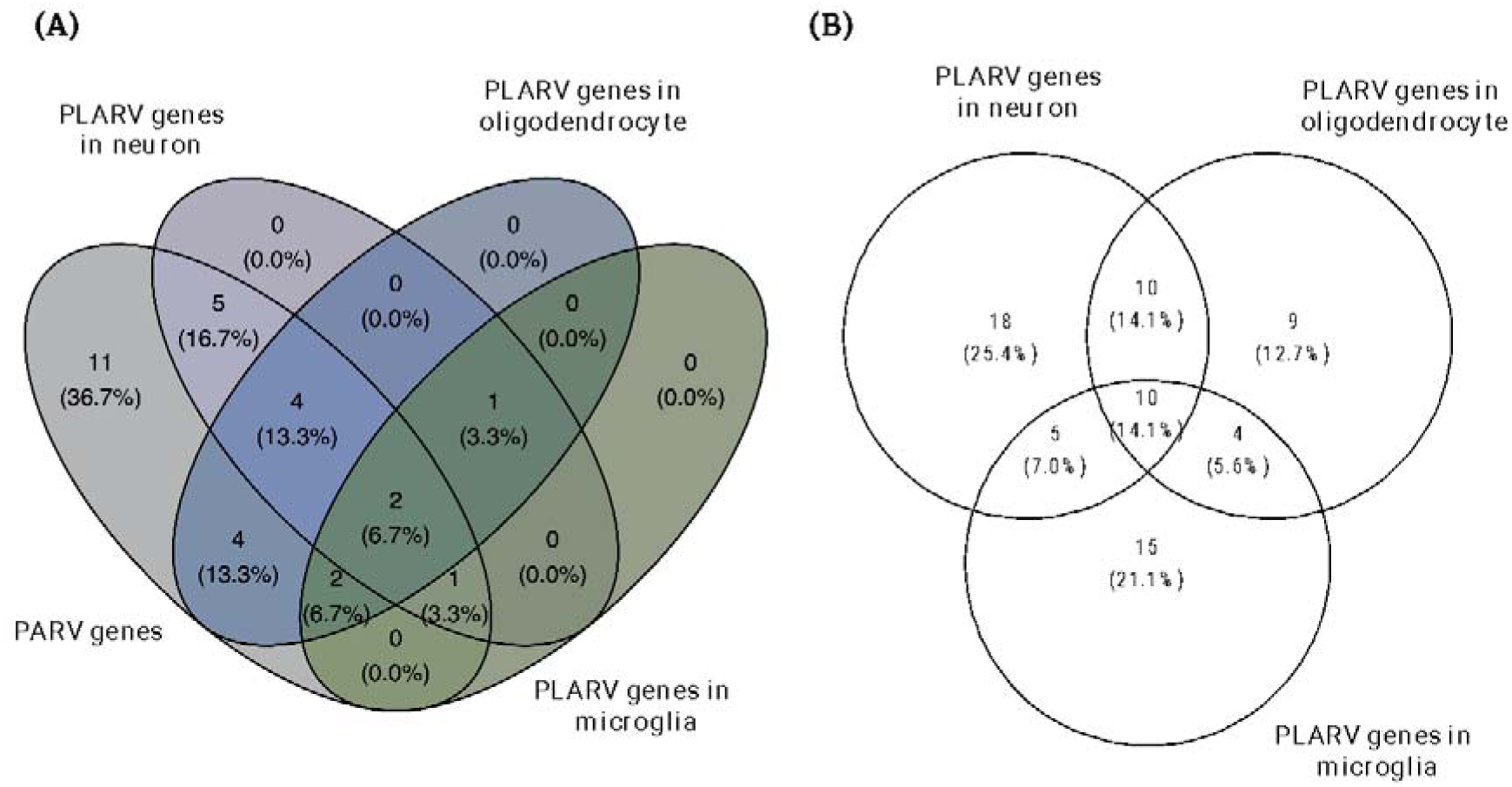
Venn diagram for *de novo* PARV and PLARV genes. (a) Variants falling inside a promoter (n = 29) are here represented in terms of the associated PARV and/or PLARV gene. Genes containing promoters harboring ASD regulatory variants are represented in the “PARV genes” category. Genes that are PLAC-linked to ASD regulatory variants are represented in the “PLARV gene” categories, one for each cell type analyzed here (neuron, oligodendrocytes, and microglia). Note that the purpose of this graph is to represent the overlap between PARV and PLARV genes, and thus, PLARV genes that do not overlap with PARV genes are here excluded. (b) PLARV genes ascribed to variants falling, or not, inside a promoter in each of the three different cell types.

We further defined PLAC interactions between active promoters and *de novo* variants and identified 69 PLARV genes linked to these variants across the three cell types. Forty-one of the total number of PLARV genes were identified in neurons, of which 18 were not detected in any other cell type (Figure 6 B).

When comparing PLARV and PARV genes, we found that in most instances (63.3%) genes with promoters harboring ASD regulatory variants were PLAC-connected to a region affecting the same gene. In other words, the gene influenced by the regulatory variant is often the “closest gene” in proximity to that variant. However, in the reimaining 36.7% of the instances they did not overlap, which means that the “closest gene” is not compulsory the variant-regulated gene (Figure 6 A).

Moreover, out of the 18 variants falling outside promoters, 14 were PLAC-linked to a promoter, and thus assigned to the corresponding PLARV gene (77.78% of the instances) (Supplementary Table 1).

This yields a total of 43 variants (91.49%, out of the final list of high regulatory impact *de novo* variants) located within promoters and/or within regulatory regions PLAC-linked to promoters, and 39 variants with at least one PLARV associated gene (82.98%).

#### 2.6.2 Inherited

From the total of 13,258 high regulatory impact ultra-rare inherited variants, 6,315 fell inside a putative promoter (47.63% of the instances), corresponding to 4,500 different PARV genes (Figure 7 A; Supplementary Table 2). In 71,18% of the instances, PARV genes were identified by a unique high regulatory impact variant inside their promoters. However, some PARV genes harbored up to 8 different variants in their promoters (namely*, RNF152* and *MIDN*).

**Figure 7.**
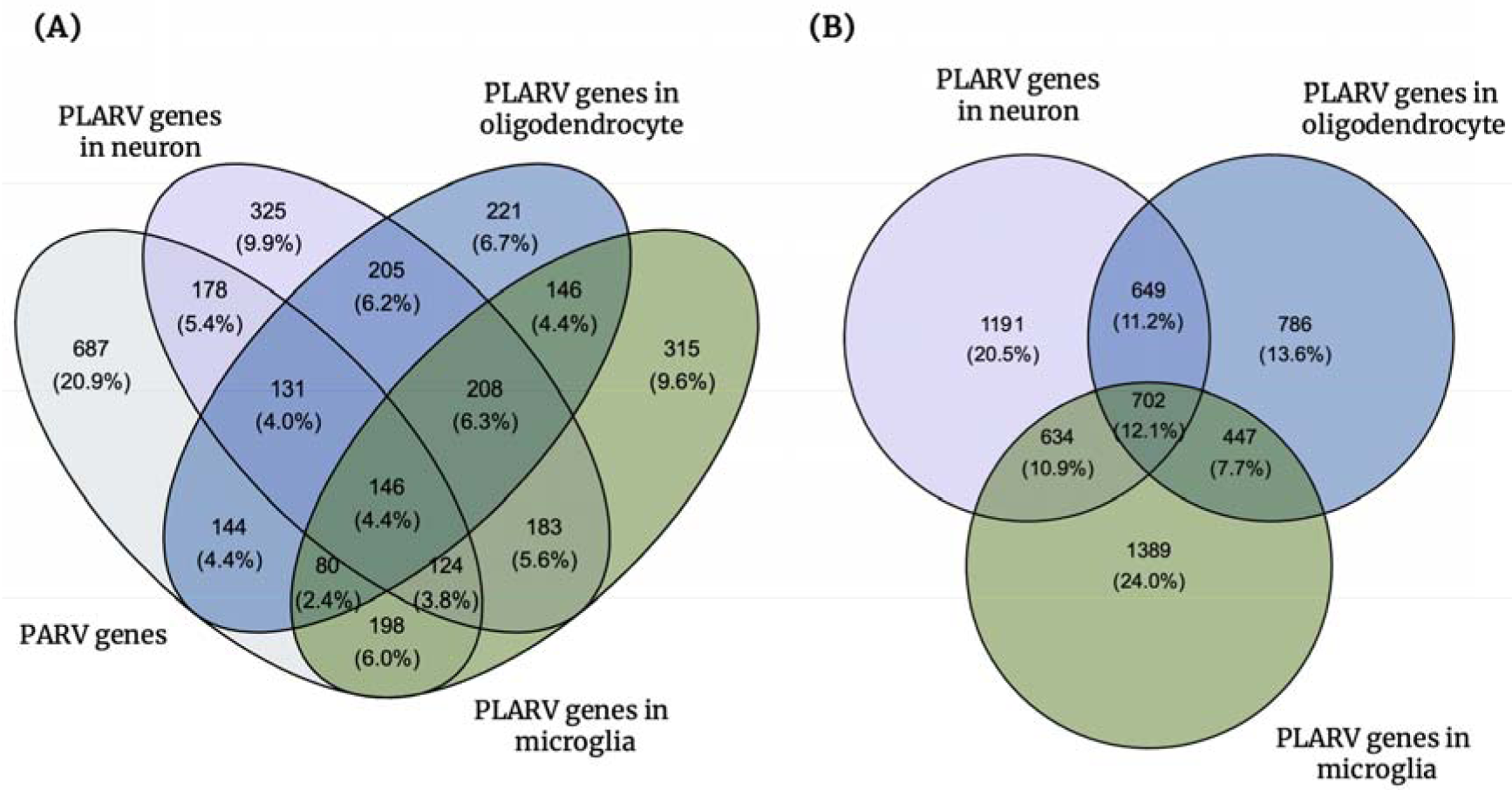
Venn diagram for inherited PARV and PLARV genes. (a) Variants falling inside a promoter (n = 6,315) are here represented in terms of the associated PARV and/or PLARV gene. (b) PLARV genes ascribed to variants falling, or not, inside a promoter in each of the three different cell types. For further explanation see caption in Figure 6.

We further defined PLAC interactions between active promoters and ultra-rare inherited variants and identified 5,798 PLARV genes linked to these variants across the three cell types. Out of the total number of PLARV genes, 3,172 were identified in microglia, of which 1,389 were not detected in any other cell type (Figure 7 B). In regard to neurons, we identified more PLARV genes (n = 3,176) but only 1,191 were neuron specific.

When comparing PLARV and PARV genes, we found that in some instances (79.10%) genes with promoters harboring ASD regulatory variants were PLAC-connected to a region influencing the same gene. However, we found that they did not overlap in 20.90% of the instances (Figure 7 A).

Moreover, out of the 6,943 variants falling outside promoters, 5,510 were PLAC-linked to a promoter in at least one cell type, and thus assigned to the corresponding PLARV gene (*i.e.*, 79.36% of the variants without a PARV gene were associated with a PLARV gene) (Supplementary Table 2).

This yields a total of 11,825 variants (89.19%, out of the final list of high regulatory impact inherited variants) located within promoters and/or within regulatory regions PLAC-linked to promoters, and 11,203 with at least one PLARV associated gene (84,50%).

#### 2.6.3 Gene function

When analyzing 68 PLARV genes associated with the final list of *de novo* high regulatory impact variants, we did not get any significant enrichment for any GO category. However, top BP (based on the gene ratio) included regulation of protein stability, dendrite development and chromatin remodeling, in line with previous evidence.

Nonetheless, when focusing on the final list of PLARV genes associated with the inherited variants, we did find significant enrichments in MF categories relevant for ASD such as ubiquitin protein ligase binding (q-value = 1.12 x 10^-9^), DNA-binding transcription factor binding (q-value = 6.51 x 10^-8^) and transcription coactivator activity (q-value = 2.21 x 10^-5^), that remained significant even when independently assessed in each cell type. The same observation was noted for CC categories like nuclear envelope (q-value = 1.13 x 10^-14^), ubiquitin ligase complex (q-value = 8.61 x 10^-9^), neuron to neuron synapse (q-value = 3.12 x 10^-8^) and spliceosomal complex (q-value = 3.19 x 10^-6^).

Furthermore, we found significant enrichments in BP categories relevant for ASD such as RNA splicing (q-value = 4.33 x 10^-10^), regulation of neuron projection development (q-value = 1.67 x 10^-7^), axonogenesis (q-value = 1.67 x 10^-7^), and histone modification (q-value = 1.52 x 10^-6^). Top enriched terms remained significant even when analyzing the three cell types separately (Figure 7).

### 2.9 TF enrichment

Given the enrichment in categories related to TF binding in the GO analysis, and the fact that the most represented sequence class in our dataset pertains to CTCF binding, we aimed to characterize further enrichment in TF binding in our final list of *de novo* and inherited variants (Supplementary Tables 4, 5). Of note, at the time this analysis was performed, the ENCODE version 2 browser was inaccessible due to the release of ENCODE version 3. Consequently, TF enrichment data was obtained from alternative sources. (see Methods).

As anticipated, CTCF demonstrated a significant association, and thus, our previous findings were corroborated.

### 2.10 Variant prioritization and diagnostic yield

Although the primary aim of this chapter was not to provide a clinical diagnosis, limited by the incapability to robustly establish a genotype-phenotype relationship in the methodology discussed here, we sought to leverage the information from variants ascribed to PLARV genes.

To avoid a huge burden of interpretation, under emerging recommendations for clinical interpretation of the non-coding genome^13^, we have focused on variants located in non-coding regions of the genome only if (i) they are within regulatory elements with functionally validated connections to target genes as proven by the Hi-C analysis here leveraged, and (ii) those genes are known to be associated with ASD risk, as compiled in our custom ASD risk gene list (Supplementary Table 6).

By following this approach, we found: one patient with a *de novo* variant PLAC-linked to *PSMD1* specifically in neuron and one patient with a *de novo* variant harbored in *POGZ*’s promoter (Supplementary Table 1). Both patients have a co-diagnosis of ID.

Moreover, we found 489 inherited variants with interactions to 124 different PLARV genes included in our custom list of ASD-associated genes. These were present in 185 different patients, with an average of 2.64 ± 1.69 variants per child (Supplementary Table 6).

Interestingly, we found one patient who harbored 10 different variants (which was the maximum observed number of variants per patient) and had one affected sibling (who has not been sequenced). All 10 variants were present in different TADs and corresponded to 10 different PLARV genes (most of them were detected specifically in neuron (Supplementary Table 6).

Additionally, one patient with a Tourette’s syndrome diagnosis harbored 9 inherited variants across various genes. Moreover, one patient harbored 8 variants, while four other patients harbored 7 variants. Among these five patients, three were diagnosed with ADHD, and one had ID. All variants harbored in each patient were present in different TADs.

## DISCUSSION

In this chapter, we have scrutinized targeted sequencing data from 200 ASD trios, where *de novo* LGD mutations and large CNVs, crucial in regulation of gene expression, had eluded detection in prior WES and microarray analyses. Moreover, we have applied the Sei framework, which is the most comprehensive chromatin-level sequence model to date. To our knowledge, this is the first time it has been applied to an ASD cohort, and we anticipate that further fruitful findings will result from larger cohorts.

Discussions about the limitations in deciphering the non-coding genome have been presented in earlier sections. Specifically, the absence of a non-coding counterpart to the triplet code, found in protein coding regions, has been a crucial obstacle for predicting the impact of mutations on gene function within the non-coding genome. Nevertheless, in this study, we have integrated data from ATAC-Seq and PLAC-Seq (which incorporates ChIP-Seq) to annotate variants based on four non-coding annotations: (i) regions of open chromatin, where DNA is exposed, thus allowing protein binding; (ii) regions of active chromatin, where epigenetic marks suggest transcription of a nearby gene; (iii) transcription factor binding sites, and (iv) prediction of the regulatory gene target using proximity to the variant and/or physical interactions genome-wide.

Importantly, in terms of general conclusions: we have identified that 28% of *de novo* variants and 25% of inherited variants exhibit a high regulatory potential in patients with negative results from WES and microarray analysis, as assessed by Sei.

By incorporating PLAC-Seq data, we were able to functionally assign a gene for ∼80%/85% of *de novo* and inherited variants, respectively. Notably, when a variant was in close genomic proximity to a PARV gene, these associations were not corroborated by functional annotations in as much as 36.7% of the instances, in line with previous findings^36^ (*i.e.,* PARV and PLARV genes did not overlap, as illustrated in Figures 6A and 7A).

Finally, while initiatives like ENCODE provide invaluable insights into defining regulatory elements of the genome, caution should be exercised when attempting to prioritize a specific type of regulatory element based on a priori hypotheses of heightened influence on gene regulation: many of the sequence classes found to be affected in ASD in this study were not significantly enriched in a specific type of cCRE signature (Supplementary Table 3).

### 3.1 Exploring the sex bias in regulatory variation whithin ASD

In our cohort, the male-to-female ratio was 4.0, consistent with previous findings^28^. This ratio persisted even when prioritizing high-impact regulatory variants, with a ratio of 3.7 within *de novo* variants and 4.03 within inherited variants.

However, in evaluating transmission bias for ultra-rare inherited variants, we detected a statistically significant transmission bias favoring inheritance from the mother that remained significant even when the analysis was limited to autosomes (Supplementary Table 2).

The debate surrounding the excess of germline DNMs arising on the maternal chromosome and the transmission bias favoring maternal inheritance has been a longstanding and essential aspect of ASD etiology^4,31,37,38^. Theoretically, if females exhibit reduced vulnerability to ASD, as evidenced by the consistently observed female-to-male ratio of 4:1 in this and most ASD studies, and individuals with ASD experience reduced fecundity, basic genetic principles predict that mothers are more likely sources of risk alleles than fathers.

Initial observations relating to the observed sex bias indicated a trend of excess maternal *de novo* CNVs, although the pattern did not achieve statistical significance. Notably, females with ASD exhibited a higher number of *de novo* CNVs compared to males, and these genomic imbalances were both larger and impacted a greater number of genes^39^. Later, some of the first statistically significant genetic evidence of a transmission bias from mothers to their sons was observed for protein-coding SNVs in the study conducted by Krumm *et al.*^40^.

On the contrary, one study identified a significant paternal-origin effect for CRE-SVs^4^. One possible explanation offered for the observation that mutations in regulatory regions follow an opposite pattern to what is predominantly seen in coding mutations, is that paternally inherited regulatory mutations, due to their less damaging impact, may require a greater number to manifest in disease. Crucially, this has given rise to an alternative hypothesis, suggesting that regulatory mutations may adhere to distinct principles compared to coding mutations.

Nevertheless, in line with our results, instances of the transmission of non-coding putative regulatory DNA from the mother to the male proband have been noted in genes such as *DSCAM* and *TRIO*, and both of them have been observed to be disrupted in ASD^1^.

Taken together, our results represent, to the best of our knowledge, the first demonstration, in a non-biased global study of the non-coding genome, that parent-of-origin effects may follow similar rules as coding variation. However, these results should be cautiously interpreted and the comprehensive contribution of a gender bias in non-coding variation to ASD needs to be determined through much larger studies. Still, this underscores that influences of sex bias on genetic risk for ASD are more intricate than previously understood, and the allelic spectrum of variants varies between the maternal and paternal genomes.

#### -Sequence class dysregulation

Probably, the most important observation in this study is the implication of a global dysregulation of CTCF.

CTCF is reported as a high confidence gene (score = 1S) in SFARI, associated with Tourette syndrome^41^, ID^42^, and, more recently, with ASD^31,37,43^. In fact, it was identified by Zhou *et al.*^44^ as a gene reaching exome-wide significance in their analysis of rare *de novo* and inherited coding variants in 42,607 ASD cases, (p < 2.5 x 10^-6^) and thus included in our list of high risk ASD genes (Supplementary Table 6).

Interestingly, in our random selection of cCREs, only 2.6% were classified as CTCF in ENCODE (Figure 2 B), and this proportion drastically decreased when we filtered for *de novo* and inherited mutations. Moreover, we observed a significant depletion of CTCF cCREs in the final list of inherited variants (Table 2).

However, when prioritizing variants with a high regulatory impact, not only did their proportion increase, but they became the regulatory class most represented amongst the detected mutations. Specifically, an increased number of mutations was associated with a reduction (and not an increment) in the CTCF regulatory score, and this difference attained statistical significance for inherited variants (p = 3.11 x 10^-8^) (Table 2). Although *de novo* variants, constrained by a limited statistical power in terms of numbers, did not reach significance, they showed a similar trend.

Furthermore, these results were corroborated from the TF enrichment analysis (Supplementary Tables 4, 5).

With this regard, it is important to note that the targeted sequencing approach followed in this chapter only covers 0.70% of the genome. Moreover, the selection of regulatory elements was performed in an arbitrary fashion, as it did not involve a priori hypotheses about which elements were most likely to influence gene expression. We solely restricted the regulatory elements to those active in tissues relevant to ASD. Thus, if we were to follow a WGS strategy, the results presented here suggest that we would likely obtain the same outcomes, supported with greater statistical power.

As further evidence, if we consider the top 10% of mutations with the highest class scores, they exclusively belong to the CTCF class, both for inherited and for *de novo* variants (Supplementary Tables 4, 5).

Moreover, CTCF was one of the most frequently disrupted TF motifs in ASD cases *versus* controls in a single-cell analysis of gene expression and chromatin accessibility^34^, and in this study. This suggested a potential mechanistic impact on the chromatin architecture: deletion of CTCF binding sites or lost on CTCF binding affinity can lead to formation of new TADs, which can cause genes to contact enhancers normally spatially too far away to do so (a process known as “enhancer hijacking”)^34^.

Of note, while writing this discussion, Nakamura *et al.*^45^, published an analysis of promoter DNV from WGS data in a cohort of 5,044 ASD patients and 4,095 siblings. Interestingly, among the findings in their analyses, they specifically identified the enrichment of ASD-gene TAD promoters at CTCF-bound regions, further corroborating our results.

In addition, Nakamura *et al.*^45^, found a specific association of DNV in TADs containing ASD risk genes and identified specific TADs with enrichment of promoter DNVs. Thus, we aimed to assess whether the variants in each of the patients with elevated numbers of high impact inherited variants were located within the same and/or adjacent TADs. If this were to be corroborated, we could hypothesize that the loss of CTCF binding affinity in TADs relevant to ASD might collectively contribute to disease risk by simultaneously affecting the expression of pertinent genes. On the contrary, we found that these variants were present in different TADs. Still, these findings prove interesting as they offer mechanistic insights into how non-coding regulatory variation may be influencing ASD risk.

Overall, it seems reasonable to assume that a substantial part of the non-coding genome in ASD is dysregulated in terms of its 3D-structure, and that further characterization of chromatin loop alterations (for example, by Hi-C analysis) will prove fruitful in further unraveling the missing heritability in ASD.

Yet, it has been reported that CTCF contributes not only to the formation of TAD boundaries but also to the stabilization of promoter-enhancer interactions. Thus, it may also be possible that TAD boundaries remain unaffected, but changes in promoter targeting by enhancers (which is influenced by CTCF-binding) are responsible for the changes in gene expression leading to disease^46,47^.

### 3.2 Biological, molecular, and cellular consequences of regulatory variants

The determination that non-coding variants predominantly target the same functional processes as protein-coding genes associated with ASD strongly supports the idea that non-coding variants may have causal roles in ASD development^12,37^. We thus sought to assess whether this holds true within our variant dataset. To do so, we conducted a burden analysis across all the gene sets listed in Gene Ontology. This analysis aimed to identify gene pathways enriched in genes affected by high-impact regulatory variants.

We found a significant enrichment of variants in pathways involved in “chromatin organization”, “RNA processing translation” and “synaptic transmission” among others, which is largely consistent with previous findings (Figure 8). The association signal for the identified genes originates from rare inherited variation. While not achieving statistical significance for *de novo* variants, our results indicate that these variants impact BP in a manner consistent with those influenced by rare inherited variation. Additionally, both *de novo* and inherited genes contribute to a shared protein-protein interaction network with a total of 1,163 interconnected protein products, enriched for expression in the central nervous system (p < 1 x 10^-6^) (data not shown).

**Figure 8.**
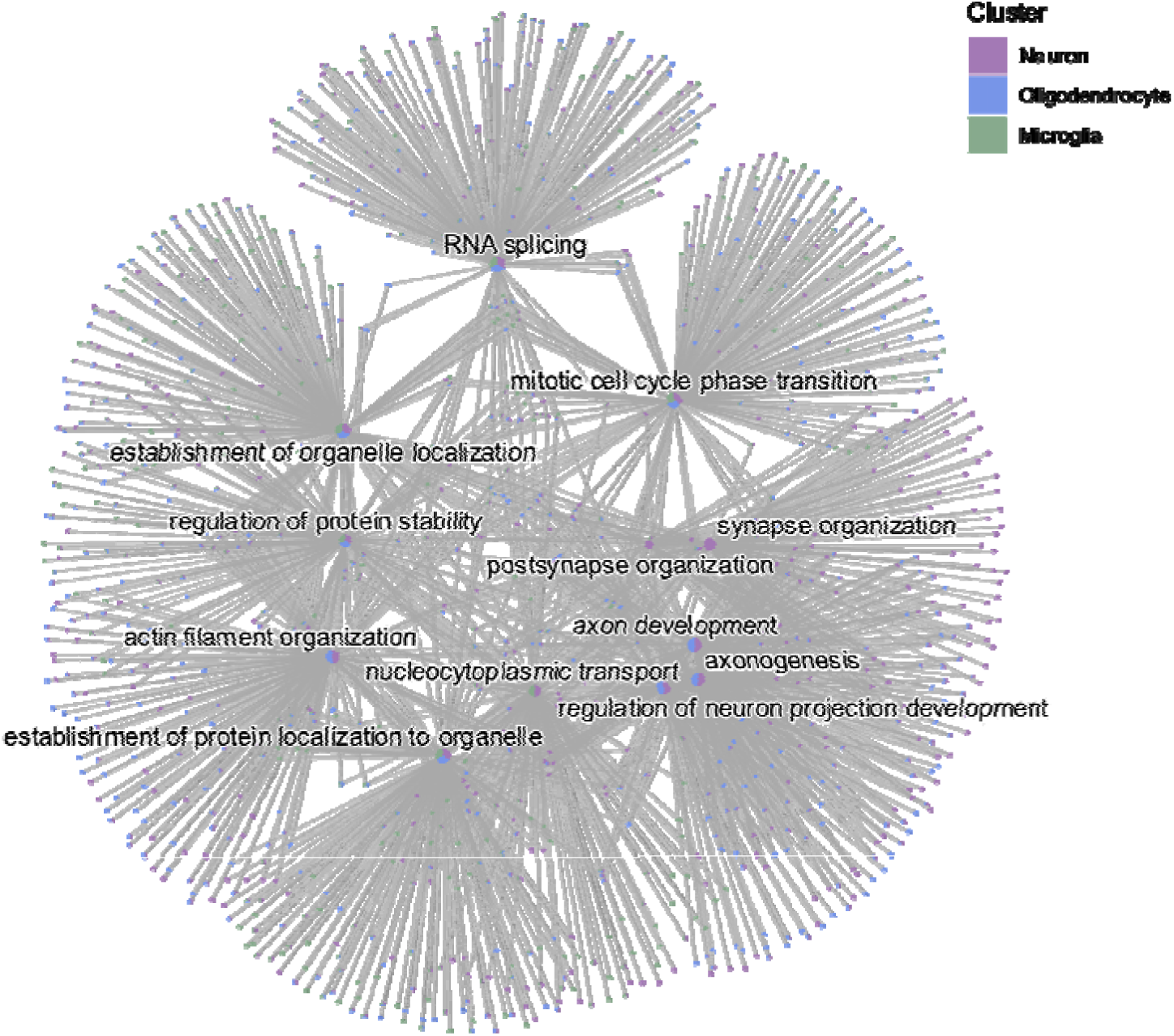
Biological processes significantly enriched within PLARV genes from the inherited variant list across the three cell types. The graphic depicts top 5 enriched terms for each gene cluster (*i.e.*, PLARV genes in each cell type). Circles represent the number of genes, with larger circles including 160 genes and smaller ones including 71.

However, the key question remains as follows: whether variation found in the non-coding genome is targeting the same set of genes that have been implicated in ASD by WES and CNV studies or a unique group of genes missed by previous efforts. When contrasted to our custom list of high-risk ASD genes, we have found that the majority of high-impact regulatory variants, whether *de novo* or inherited, are PLAC-linked to genes not previously associated with ASD. Still, GO enrichments point to a scenario where coding and non-coding variation synergize in already characterized ASD relevant pathways. This aligns with recent observations indicating that rare inherited variation predominantly affects previously unknown risk genes impacting similar pathways to those influenced by known genes^22,26,44,48^.

Overall, this convergence supports the idea of a causal contribution of non-coding regulatory mutations to ASD etiology.

### 3.3 Preliminary genotype-phenotype associations

Although it was not the primary aim of this study, we find it beneficial to highlight the following scenarios in regard to theoretical genotype-phenotype correlations. Nonetheless, we acknowledge limitations in our conclusions, as functional characterization of the evidence discussed in this section will be necessary to robustly establish evident interactions between non-coding variation and phenotype consequences.

Firstly, when analyzing high-impact inherited regulatory variations, and before prioritizing variants associated with PLARV genes already established to have a functional link to ASD, we identified two genes, *RNF152* and *MIDN*, each with eight different variants (Supplementary Table 2). Moreover, all these variants were also PLAC-linked to the corresponding gene, and we further characterize 3 variants outside *MIDN’s* promoter but PLAC-linked to the gene.

*RNF152* has been characterized to catalyze the ubiquitination of Rheb, a small GTPase enriched on the lysosomal membrane, where it activates mTORC signaling^49^. The ubiquitination of Rheb suppresses mTORC1 activation, which, in turn, has been found to be aberrantly activated in ASD patients^50^. In this regard, we hypothesize that the elevated number of regulatory variants, collectively affecting *RNF152* gene expression, leads to reduced levels of its protein product, thereby resulting in overactivation of the mTOR signaling pathway.

*MIDN*, on the other hand, has been linked to Parkinson’s disease and regulates neurite outgrowth^51^. Moreover, it has been associated with female ASD in a WES^52^. Although most of our variants are found in male patients, these evidence points to a probable true association with ASD.

While these two genes are not currently listed in the SFARI database, we present additional evidence supporting their potential implications in ASD etiology.

Furthermore, several lines of evidence point to more severe phenotypes relating to higher burdens of genomic load. In particular, earlier studies have identified an inverse correlation between the IQ of individuals with ASD and the burden of specific regulatory variants^12^.

In this light, it is interesting that our analysis uncovered a patient with 10 distinct high-impact regulatory variants with PLARV genes included in our list of ASD-risk genes (Supplementary Table 6). This individual stands out as the sole case within our sample cohort who has a documented sibling with ASD. Considering that multiplex families often carry an elevated burden of inherited mutations and given the negative results for both exome and microarray analyses in this family, we propose that there is a slight possibility that the cumulative effects of the high-risk inherited variants described here are influencing disease risk and contributing to the observed recurrence in this family.

However, cautiousness should be taken when interpreting these results. First, it would be ideal to confirm, by sequencing methodologies, that these variants are also present in the affected sibling, and they are absent in a suitable control cohort. If so, functional assays should be performed in order to properly define their mode of action. Given that they are situated within different TADs, the likelihood of disrupted promoter-enhancer interactions within a specific genomic region, as observed elsewhere^45^, and thus the possibility of them collectively functioning as a mutational hotspot, is ruled out.

Moreover, the second patient with the highest number of variants in PLARV genes which are known ASD-risk genes, harbors two different variants PLAC-linked to *FAM98C* (Appendix 10.2, Supplementary Table 10). *FAM98C* is classified with SFARI score 2. Initially, the identification of a PTV in the gene linked it to ASD^53^. Furthermore, a recent TADA analysis definitively identified *FAM98C* as an ASD candidate gene with a FDR < 0.1^6^. Our analysis provides further evidence to associate the gene with disease risk.

### 3.4 Limitations and future perspectives

The primary limitation in this study is the absence of a sibling/control cohort. Future perspectives involve retrieving additional data from a control cohort to address this. Ideally, access to larger cohorts of ASD patients would enhance statistical power, allowing for the expansion of current conclusions drawn from inherited variants to DNVs, which have been limited in number.

Moreover, the main conclusions gathered were made possible thanks to the utilization of PLAC-Seq data from key brain cell subpopulations. However, our initial selection of cCREs was based on activity in the brain, as well as in GI tissue. While we could not draw any conclusions regarding the latter, major goals will be to extend these approaches to all diseased tissues when data becomes accessible.

While we were working on this research, Shin et al, 2024 study was published in Cell Genomics, representing a significant development in the field of non coding genome in ASD^54^. Findings are particularly noteworthy despite the differences in cohort and research design between both studies. The have focused on evolutionary signatures of selection in humans, such as Human Accelerated Regions (HARs) that are emerging as potentially significant in ASD. HARs are mostly found in non-coding regions, and research has shown that rare, inherited mutations in HARs are more prevalent in individuals with ASD, a finding supported by our analysis of active regulatory elements from the ENCODE database.

In the future, there are several important areas that require more research:i) Increasing the scope of non-coding genome studies with larger selection of active regulatory regions from *in vivo* and *in vitro* specific functional studies to enhance statistical power and applicability of results, ii) Advancing the development of multi-omic bioinfomatic tools to predict the functional consequences of non-coding variations from larger WGS studies, iv) Exploration of how non-coding variants may contribute to the heterogeneity of ASD phenotypes. In conclusion, our study contributes to the growing body of evidence suggesting that non-coding regions play a crucial role in ASD genetics. By adopting an unbiased, tissue-specific approach and leveraging advanced computational methods, we have provided new insights into the complex genetic architecture of ASD. As the field continues to evolve, the study of non-coding variants is becoming an increasingly field of ASD research, potentially leading in the near future, to improved diagnostic and therapeutic strategies for individuals with this complex neurodevelopmental disorder.

## Data Availability

All data produced in the present study are available upon reasonable request to the authors
All data produced in the present work are contained in the manuscript

## ACKNOWLEDGEMENTS

We thank the project: Instituto de Salud Carlos III (ISCIII)/PI1900809/Cofinanciado FEDER. SDA was supported by a Xunta de Galicia predoctoral fellowship.

## AUTHOR CONTRIBUTIONS

SDA wrote the paper and did all the genetic analysis.J RPT contribute to obtain raw sequencing data from the sequencing hub. CA, MP, JGP and AC contribute to the sample selection and collection. AC and CRF critically revised the work and approved the final content. SDA, CRF, AC participated in the design and coordination of this study.

## ADDITIONAL INFORMATION

Supplementary information The online version contains supplementary material available at https://

Correspondence and requests for materials should be addressed to C. RodriguezFontenla.

Reprints and permission information is available at http:/

